# The Role of Bone Marrow Microenvironment in Osteogenesis Imperfecta: Evidence from Single-Cell RNA Sequencing

**DOI:** 10.64898/2026.08.21.26361022

**Authors:** Zhiming Wu, Suzanne den Haan, Wouter Nijhuis, Claudia Y. Janda, Thanasis Margaritis, Harrie Weinans, Ralph Sakkers, Anne J. Spaans, Kelly Warmink

## Abstract

**INTRODUCTION:** Osteogenesis imperfecta (OI) is a genetic disorder primarily due to mutations in collagen type I-encoding genes, resulting in fragile bones, frequent fractures, pain, and mobility issues. Disease severity and phenotype vary widely, even with the same mutation, suggesting the importance of other factors within the bone microenvironment that influence disease severity. To study the role of such factors, we analyzed bone samples from OI patients and healthy controls using single-cell RNA sequencing to reveal if RNA expression profiles may uncover mechanisms behind OI phenotype.

**METHODS:** Bone samples from surgeries of OI patients and healthy individuals isolated and RNA single-cell sequencing was performed, followed by quality control and bioinformatics analysis. Two healthy and three OI patients were included: two with type-I OI, characterized by a mutation in COL1A1 (collagen type I), and another with type-VIII OI, associated with LEPRE1 mutations, which disrupt the 3-hydroxylation of type I collagen.

**RESULTS:** Clustering and differential expression analysis showed distinct subpopulations in mesenchymal and immune cells. In all OI samples, mesenchymal stromal cell (MSC) proportions were reduced compared to healthy controls. OI type-I patients showed decreased osteoblast numbers alongside an increase in osteoclast precursor cells. Whereas in OI type-VIII, all bone turnover-related cells (osteoblast, osteoclast precursor, and osteoclast) were elevated. Notably, BMP5 and RUNX1 were downregulated in MSCs from both OI types.

**DISCUSSION:** This study demonstrates that the bone marrow microenvironment in OI is significantly altered beyond the known collagen defects. Single-cell RNA sequencing revealed reduced MSC numbers and downregulated osteogenic gene expression. Furthermore, alterations are patient-specific: OI type-I is characterized by reduced osteoblast counts, whereas OI type-VIII exhibits increased osteoblasts and osteoclasts. These findings highlight the critical role of impaired osteogenic differentiation and an abnormal bone remodeling environment in the pathology of OI.

## Introduction

Healthy bone formation is a dynamic process involving the coordination of various cells and molecules within the bone marrow microenvironment. Osteoblasts and osteoclasts play crucial roles in bone deposition and resorption, respectively. The osteoblasts produce collagen type I, which is the primary structural protein in bone and provides a strong scaffold for mineral deposition. Collagen type I interacts with numerous extracellular matrix molecules, which together regulate mineralization, bone strength, and toughness. In the bone marrow, other resident cells such as mesenchymal stromal cells (MSCs), immune cells, and hematopoietic stem cells support this intricate process, contributing to bone homeostasis.

In osteogenesis imperfecta (OI), commonly known as brittle bone disease, these processes are disrupted, predominantly due to mutations in type I collagen or proteins responsible for or related to its posttranslational modification (1–3). The resulting altered bone matrix formation leads to reduced bone mass, frequent fractures, and skeletal deformities(1, 4). In addition to skeletal manifestations, OI patients may present with craniofacial abnormalities, joint hypermobility, hearing loss, and respiratory complications(5). In OI approximately 85% of mutations occur in the COL1A1 or COL1A2 genes(6). The genetic abnormalities in OI can decrease the quantity of type I collagen (quantitative defects) or disrupt its structure (qualitative defects), leading to a spectrum of phenotypes ranging from mild to severe(7). A glycine substitution in the triple-helical domain of type I collagen is the most common variant in classical OI and is typically associated with severe OI(8). However, a study in 3152 patients showed that specific site variants, particularly in COL1A2, are predominantly linked to lethal phenotypes (8). Lethality further increases when variants affect regions critical for extracellular matrix interactions (5). There are also individuals with the same glycine substitution showing different phenotypes(5) . Thus, although the type and location of the collagen variant contribute to phenotypic variability, additional factors such as interactions between collagen and other components in the bone marrow microenvironment are likely to modulate the clinical phenotype(3).

We hypothesize that in OI, the defective collagen often interacts with other molecules and cells within the bone environment, together leading to an altered and diminished bone matrix(9). During bone formation, collagen interacts with various proteins, growth factors, and cell types, such as integrins, proteoglycans, and MSCs, which contribute to matrix organization, mineralization, and bone strength(10). Variations in gene expression of these interacting cells and molecules may cause different bone formation processes, potentially explaining the observed phenotypic diversity in OI patients. Understanding how collagen interacts with the bone marrow cells and how these interactions differ between patients could shed light on the mechanisms underlying OI phenotypes.

To explore this hypothesis, we employed single cell RNA-sequencing (scRNA-seq), a powerful tool for studying cellular heterogeneity and gene expression profiles at the individual cell level. By analyzing bone tissue from pediatric OI patients and healthy children undergoing bone surgery, we aimed to uncover differences in cell populations and gene expression profiles within the bone microenvironment that may underlie disease severity. This approach offers new insights into the complex molecular landscape of OI and may help identify therapeutic targets to improve patient outcomes.

## Methods

### Sample Collection

To gain insight into the cellular landscape of bone tissue, we employed scRNA-seq on bone-derived cells from both healthy individuals and OI patients. Tissue samples were collected from human surgery waste, following national and institutional guidelines, with informed consent from biobank number 18-685 and tissue release protocol number 26-041. Bone fragments and bone marrow from 5 children were collected. Two otherwise healthy children undergoing surgery for knee joint epiphysiodesis and precise nail right tibia and osteotomy right fibula. Three OI patients, undergoing surgery for placing 8-plate, reconstructive surgery of the hip, and placing nail right femur, of which two patients were OI type-I and one patient was OI type-VIII. Collected material was freshly processed for single cell isolation (Figure 1A).

**Figure 1:**
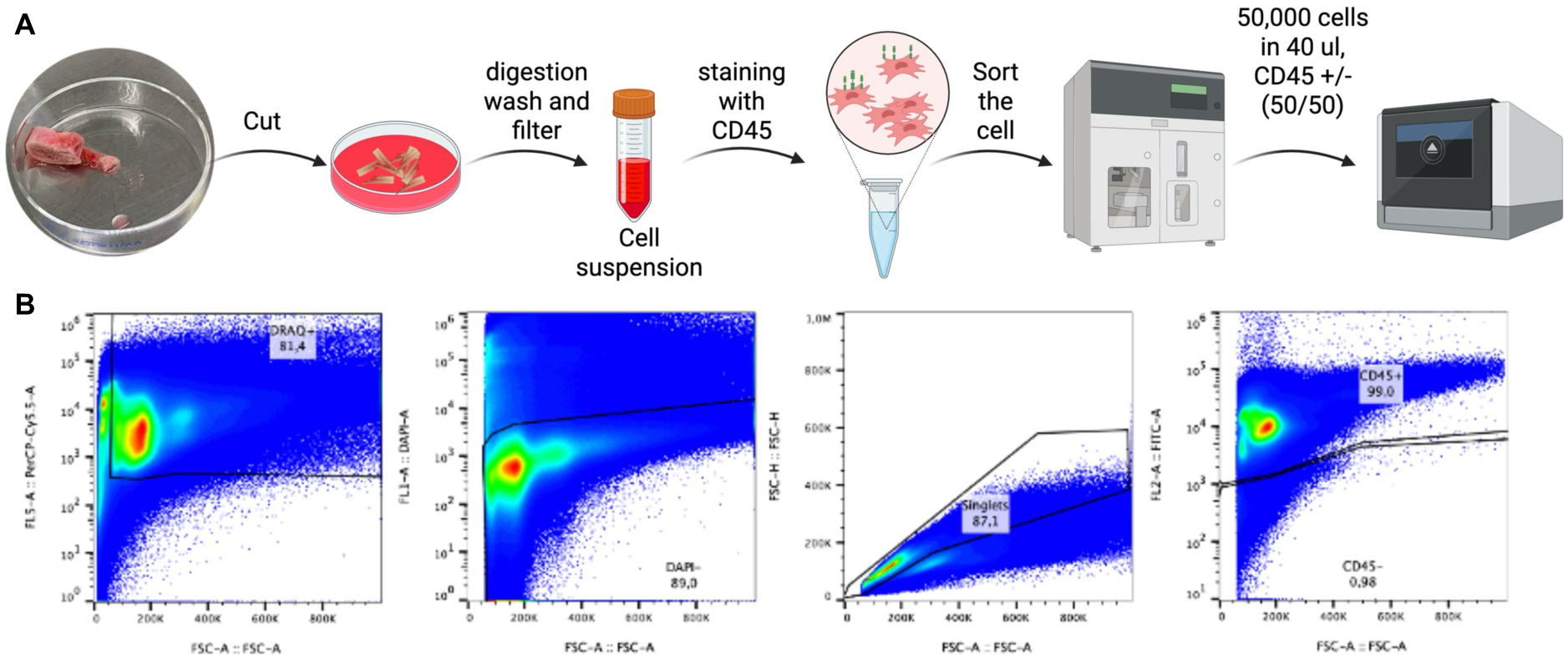
Overview of scRNA-seq and flow cytometry graphs. **(A)** Schematic representation of the workflow used for scRNA-seq analysis, where single cells were isolated, stained with CD45 and sorted to enrich non-immune cell populations to 50%. **(B)** Patient-derived cells were stained with DRAQ5, DAPI, and CD45-FITC antibodies. Cells were pre-gated on DRAQ+, DAPI-, singlets, CD45+, and CD45- cells. The percentage of gated cells within the population is indicated. The data represent samples from 5 different patients.

### Single cell isolation

Single cells were isolated by mechanical dissociation followed by two rounds of enzymatic digestion performed for 30 minutes at 37 °C using 0.1 mg/mL Liberase™ DL (Sigma-Aldrich, Cat. No. 5466202001) for the first digestion and 0.1 mg/mL Liberase™ TM (Sigma-Aldrich, Cat. No. 05401119001) for the second digestion. After each digestion step, bone fragments were washed with phosphate-buffered saline (PBS) and filtered through a 70 μm cell strainer. The collected cells were centrifuged and resuspended in fluorescence-activated cell sorting (FACS) buffer containing 1% bovine serum albumin (BSA), 1 mM ethylenediaminetetraacetic acid (EDTA), and 10 μM Rho-associated protein kinase (ROCK) inhibitor Y-27632 (Biogems, Cat. No. 1293823). Red blood cells were lysed using red blood cell lysis buffer (Invitrogen) for 4 minutes at room temperature. Subsequently, cells were incubated with 20 μg/mL of fluorescein isothiocyanate (FITC)-conjugated cluster of differentiation 45 (CD45) antibody (Stemcell, Cat. No. 60161.2, Clone 2.462) for 30 minutes at 4 °C in the dark. Live and dead cells were discriminated using 5 μM DRAQ5 (a far-red DNA-binding dye; BioLegend) and 2.5 μM 4ʹ,6-diamidino-2-phenylindole (DAPI; BioLegend).

Mesenchymal stromal cells (MSCs) and their osteoblastic progeny are relatively sparse compared with immune lineage cells in bone marrow. To obtain a balanced representation of mesenchymal lineage cells (CD45-negative) and immune lineage cells (CD45-positive) within bone and bone marrow samples, cells were sorted into CD45-positive and CD45- negative populations using a Sony SH800 cell sorter equipped with a 100 μm nozzle chip.

Sorting was performed by selecting DRAQ5-positive, DAPI-negative, single cells. Droplet calibration was carried out using Automatic Setup Beads (Sony, Cat. No. LE-83001). Following sorting, CD45-positive and CD45-negative cells were mixed at a 50/50 ratio, and 45,000 cells per condition were used for droplet-based single-cell RNA sequencing (scRNA-seq) using the 10x Genomics Chromium single-cell platform. Cells were loaded according to the manufacturer’s protocol for the Chromium Single Cell 3ʹ Gene Expression Kit. Libraries were prepared using the 10x Genomics Chromium 3ʹ Gene Expression Solution v3.1 and sequenced on an Illumina NovaSeq 6000 sequencing system. An overview of the experimental setup is provided in Figure 1.

### scRNA-seq data processing

Sequencing data were demultiplexed and converted to FASTQ format using the mkfastq function from the CellRanger toolkit (v6.1.1). Reads were aligned to the human GRCh38_3.0.0 transcriptome, and feature count matrices were generated with the count function in CellRanger.

A raw Seurat object was created in R (v4.4.0) using the read10Xlibs function from the scUtils package (v1.123). Mitochondrial gene expression was assessed to determine mitochondrial content, while hemoglobin gene expression was analyzed to estimate erythroid contamination. Cells were filtered out if they contained fewer than 1,000 unique transcripts, more than 15,000 unique transcripts, over 10% mitochondrial transcripts, or more than 3% hemoglobin transcripts. Doublets were annotated using ScDblFinder (v1.10) on the RNA assay and data slots of the Seurat object(11) (nfeatures = 1,000, iter = 4), and cells with a doublet probability exceeding 0.5 were removed.

The dataset was normalized using the SCTransform method(12) and analyzed with Seurat (v5.1.0)(13). The top 3,000 most variable genes were selected, and the cell cycle phase of each cell was determined using the CellCycleScoring function implemented in Seurat, using the built-in gene lists. In addition, cell cycle phase correlated genes were determined using metadataCorrelations and derivedCellcycleGenes from the SCutils package. All cell cycle phase and cell cycle correlated genes were excluded from the variable features, in addition to those related to hemoglobin, ribosomal, and stress response, yielding 2,889 variable features. UMAP was generated using the top 40 principal components (n.neighbors = 30, min.dist = 0.3). Clustering was performed using FindNeighbors and FindClusters, with 41 principal components and a resolution of 2.0. After initial PCA reduction, data was integrated to correct for donor-specific batch effects by using Seurat’s IntegrateLayers and method=HarmonyIntegration, which is a batch effect correction method iteratively optimizing to align the same cell types across different donors/batches, thereby better revealing true biological variation. Layers were rejoined after integration, and Harmony reduction was used for further UMAP visualization. The resulting 41 clusters were annotated using SingleR 2.6.0(14), with ‘HumanPrimaryCellAtlasData’ reference dataset, with the label set to main cell types. The top 5 most variable genes per cluster were used to check and refine the cell annotations by comparing to known markers from literature.

### scRNA-seq data analysis

Next, differentially expressed (DE) genes were identified per cluster and condition with the function FindMarkers from Seurat, using logfc.threshold = log2(1.5) and min.cells.group = 20. DE genes were filtered on a p.val.adj < 0.05. Top 25 DE genes were submitted to GO overrepresentation testing (Biological Process, Molecular Function, and Cellular Component) using enrichGO from clusterProfiler (v4.12.1)(15) to determine gene set enrichment. DE genes for cell clusters of interest were plotted using EnhancedVolcano (v1.22.0), while overrepresentation results for cell clusters of interest were plotted using clusterProfiler::dotplot (v4.12.1).

## Data Availability

The raw and processed scRNA-seq data will be deposited in the Gene Expression Omnibus (GEO) upon submission of the manuscript to a peer-reviewed journal. The data will be made publicly available upon publication.

## Results

### Single-cell RNA sequencing reveals diverse bone cell populations

Clustering of gene expression data from five donors (three OI, two healthy) revealed a total of 41 distinct cell clusters (0 to 40), as visualized by UMAP (Figure 2A and 2B). These clusters represent a variety of cell types, with a mixture of immune and non-immune cells. Among immune cells, we identified various populations including CD4+ T cells, CD8+ T cells, NK cells, B cells, osteoclasts, and dendritic cells. Osteoclasts and macrophages share a common origin from the monocyte-macrophage lineage and exhibit significant developmental and functional overlap. Macrophage precursors differentiate into multinucleated osteoclasts under RANKL (Receptor Activator of Nuclear Factor κB Ligand) and M-CSF (Macrophage Colony-Stimulating Factor) stimulation, mediating bone resorption (16). The UMAP also revealed distinct non-immune cell types critical to bone biology, such as osteoblasts, fibroblasts, and their MSC precursors. Cluster annotation was based on both SingleR using the HumanPrimaryCellAtlasData data set, as well as on well-established gene markers (Figure 2B), especially for the non-immune cell types that are less well established in SingleR (Supplement figure 2). CD45- cells encompassed critical components of the bone matrix such as osteoblasts (marked by *ALPL* and *RUNX2* expression), MSC-like cells, and fibroblasts.

**Figure 2:**
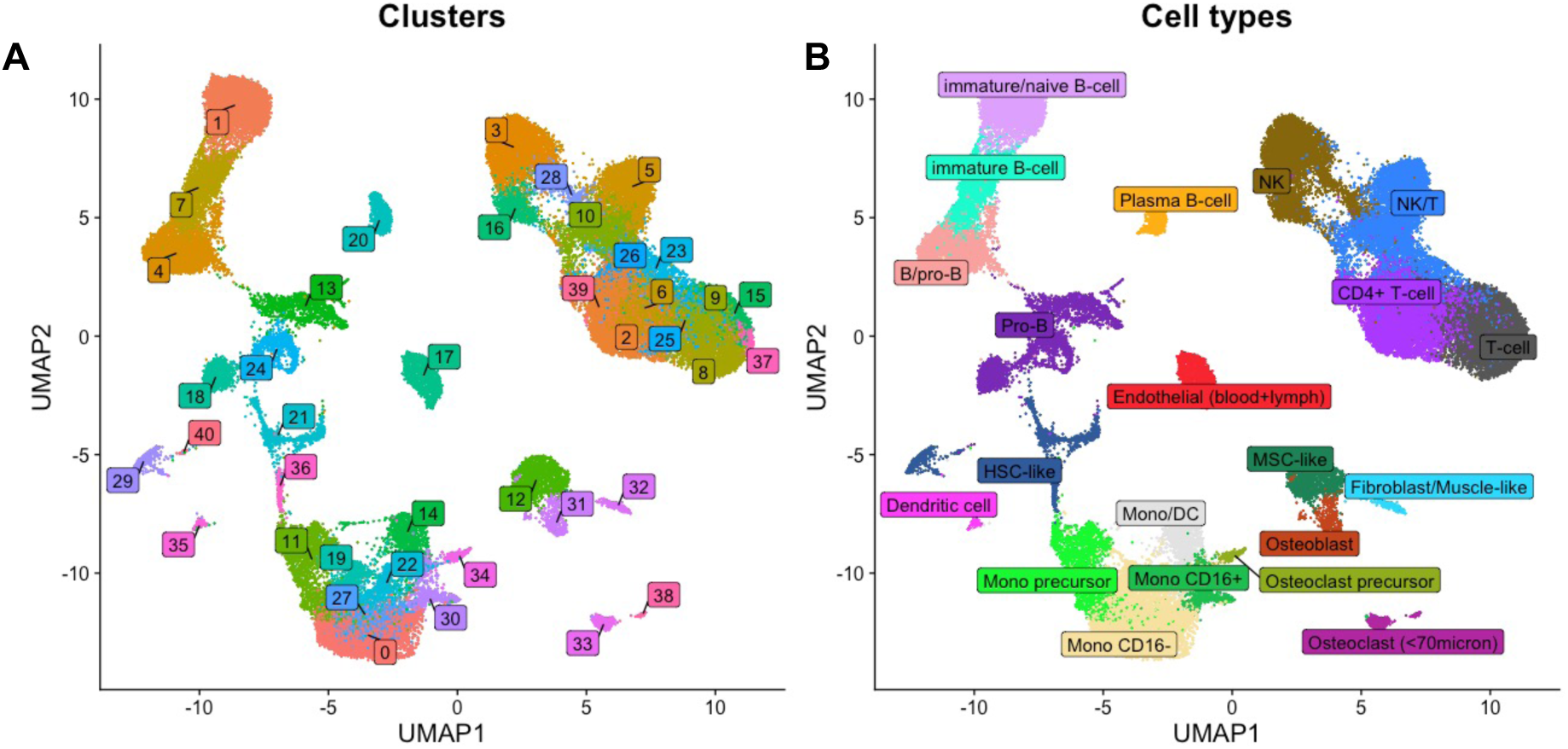
Overview of scRNA-seq and UMAP clustering of bone and immune cells. **(A)** UMAP plot showing all cells of all donors (healthy and OI), where each single cell is represented with a dot. Cells of similar RNA expression cluster together and form a total 41 distinct clusters of cells (0 to 40). **(B)** Based on gene expression profiles cell clusters are annotated with major cell types. The resulting 41 clusters were annotated using SingleR 2.6.020, with ‘HumanPrimaryCellAtlasData’ reference dataset, with the label set to main cell types. The top 5 most variable genes per cluster were used to check and refine the cell annotations by comparing to known markers from literature.

### Altered bone cell composition in OI patients compared to healthy individuals

To further explore the role of bone-specific cells in OI, we zoomed in on the bone-related cell populations (Figure 3A). In samples named ‘healthy A’ and ‘OI B’, few bone-type cells were detected. Nevertheless, our analysis identified key bone cell types in all other samples, including MSC-like cells (1.4%), fibroblast/muscle-like cells (1.5%), osteoblasts (2.6%), osteoclasts (3.4%), and osteoclast precursors (1.5%). Quantitative comparisons of cell composition between healthy and OI patients (Types I and VIII) revealed several shifts (Figure 3B, 3C; Supplement figure 1). In healthy bone, MSC-like cells constituted 4.41% of bone cells. Both OI groups showed lower MSC proportions (OI I 1.51%, OI VIII 1.39%). In addition, osteoblasts were significantly reduced only in OI type I (Figure 3B, 3C). In contrast, OI type VIII showed elevated numbers of osteoblast, fibroblast/muscle-like, osteoclast precursor, and osteoclast cells, suggesting hyperactivation of bone turnover pathways.

**Figure 3:**
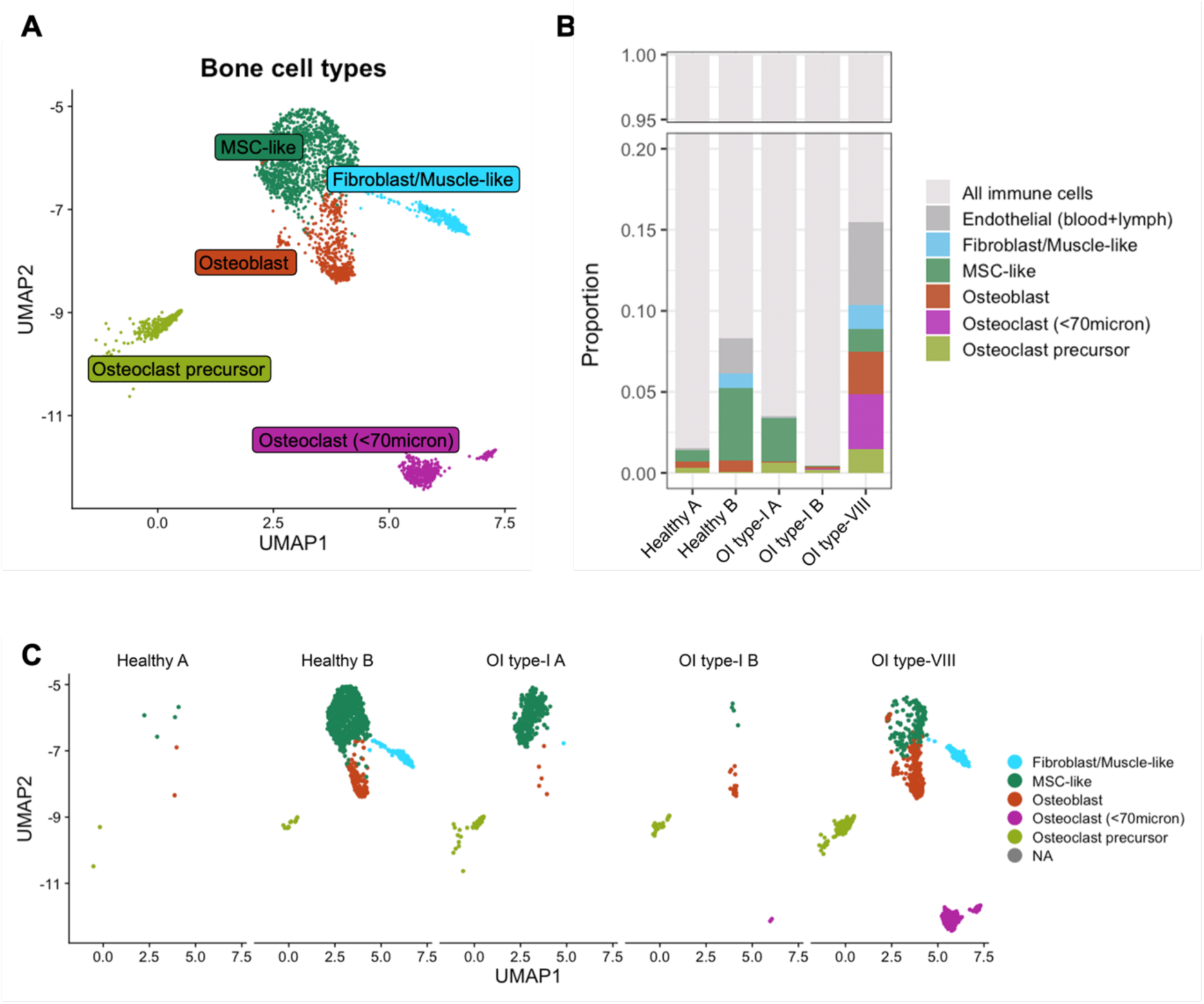
Bone cell composition differences between healthy and OI patients. **(A)** UMAP plot showing bone-related cell types (MSC-like, fibroblast/muscle-like cells, osteoblasts, osteoclasts, osteoclast precursors) across all samples. **(B)** Bar plot bone cell type proportions in healthy controls (A, B), OI type I (A, B), and OI type-VIII patients. Bone cell types are color-coded; endothelial and immune cells are grey. **(C)** UMAP projections of bone cell populations in healthy (A, B), OI type-I (A, B), and OI type-VIII patients.

### Differential gene expression and pathway enrichment in MSCs in OI patients

MSCs and osteoblast were further studied by differential gene expression analysis and Gene Set Enrichment Analysis (GSEA). GSEA analysis showed significant changes in pathways related to extracellular matrix organization and connective tissue development in OI types I and VIII compared to healthy MSCs (Figure 4A). In OI type-VIII, pathways related to cartilage development and Wnt signalling showed significant changes from healthy MSCs (Figure 4A). Furthermore, in MSCs several genes essential for osteogenic differentiation and bone signaling were downregulated in OI type-I and VIII, including RUNX1, which is critical for early osteoblast differentiation, and BMP5, which promotes bone formation (Figure 4C).

**Figure 4:**
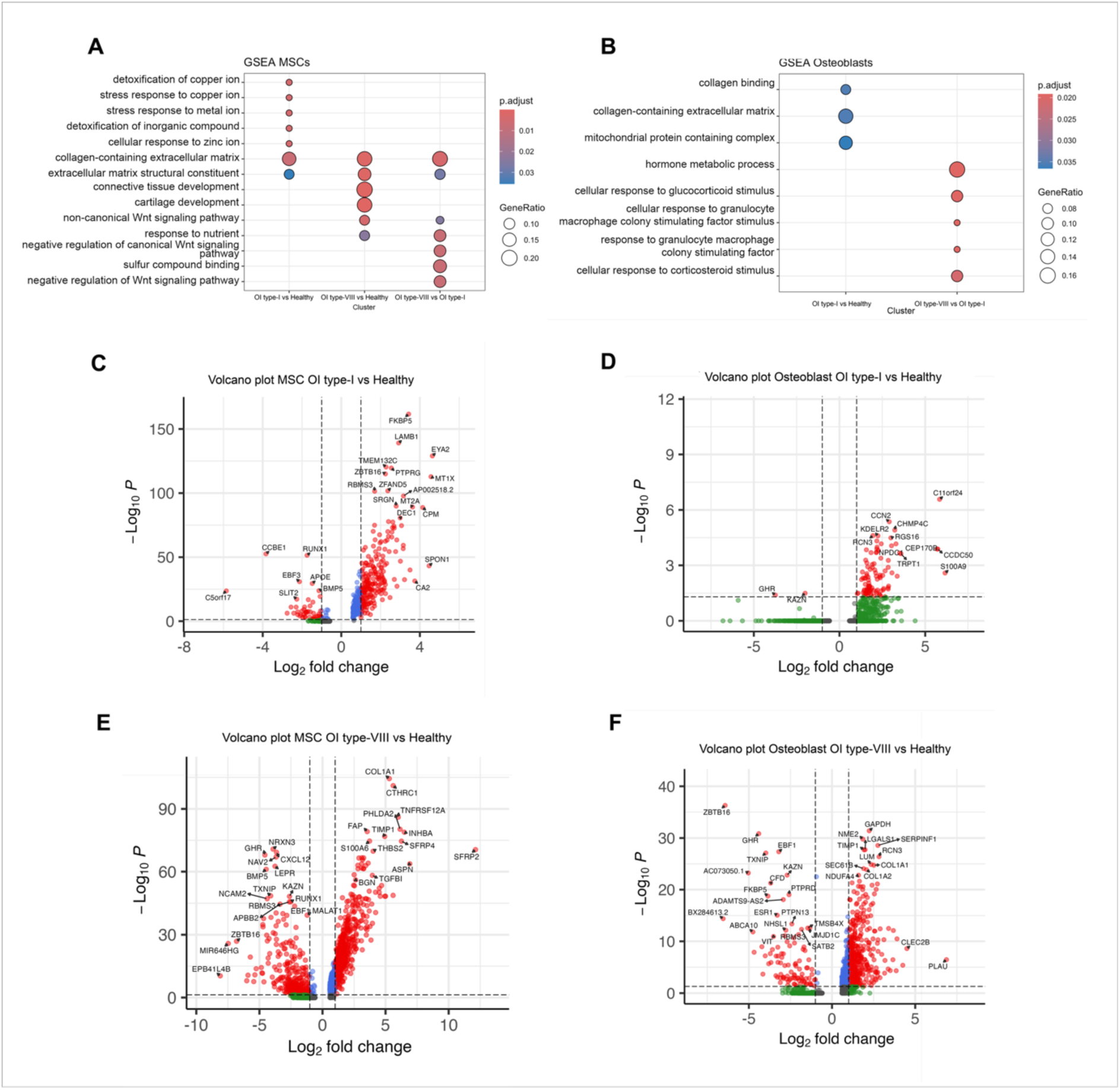
Differential gene expression in MSCs and osteoblasts (A,B) Gene Set Enrichment Analysis (GSEA) results for MSCs **(A)** and osteoblasts **(B)** comparing OI patients (Type-I and VIII) to healthy controls. **(C-F)** Volcano plots depicting differentially expressed genes in MSCs (C, E) and osteoblasts (D, F) from OI patients (type-I and type-VIII) were compared to healthy controls. Volcano plots show magnitude (log₂ fold change) and significance (-log₁₀ p-value) of gene expression differences between groups.

Additionally, genes related to extracellular matrix formation and cellular stress responses were upregulated in OI type-I MSCs. For instance, CA2, which regulates pH during bone resorption(17). (Figure 4C). Stress-related genes(18, 19), including FKBP5 and MT2A, were upregulated in OI type-I MSCs. In addition, genes implicated in cell fate regulation and differentiation, such as PTPRG and the osteogenic regulator ZBTB16, were also increased in OI type I MSCs (Figure 4C).

In addition to downregulated RUNX1 and BMP5, MSCs from OI type-VIII patients showed reduced expression of the hormonal receptors(20, 21) LEPR and GHR, key regulators of adipogenic-osteogenic balance as well as metabolic and growth-related signaling (Figure 4E). Extracellular matrix-related genes COL1A1, TGFBI, THBS2, and TIMP1 were upregulated in OI type-VIII compared to healthy MSCs, indicating increased matrix organization/remodeling and inhibited degradation (Figure 4E). However, despite increased matrix formation, the matrix produced is likely abnormal or dysfunctional due to defects in CRTAP/P3H1- associated collagen modification pathways, which underlie the severe phenotype observed in this OI subtype.

### Differential gene expression and pathway enrichment in osteoblasts in OI patients

GSEA in osteoblasts showed pathways linked to bone matrix production were altered in OI type-I compared to healthy osteoblasts (Figure 4B). In OI type-VIII compared to OI type-I, pathways related to glucocorticoid response and hormone metabolism were altered (Figure 4B). Next, we looked at genes differing between osteoblasts of OI type-I compared to healthy controls (Figure 4D). We observed reduced GHR expression in OI type-I osteoblasts, suggesting these osteoblasts may have impaired growth hormone signaling that leads to skeletal maldevelopment (Figure 4D). In contrast, inflammatory stress response and matrix regulation genes, such as S100A9(22), were elevated. Higher KDELR2 levels further point to disturbed intracellular trafficking of collagen chaperones, potentially affecting collagen folding and fibril formation through altered HSP47 recycling(23, 24) in OI type-I osteoblasts.

The gene expression profile of osteoblasts from the OI type-VIII patient (Figure 4F) also shows downregulation of GHR and KAZN compared to healthy controls, similar to OI type-I. Furthermore, reduced TXNIP suggests altered redox and metabolic regulation(25), while downregulation of ESR1 and PTPN13 points to disrupted estrogen signaling(26) and β- catenin–associated pathways(27), respectively. In addition, we observed upregulated COL1A1 and COL1A2 gene expression, as well as other matrix-related genes(28) like TIMP1, LUM (related to collagen fibrillogenesis(29)), and SERPINF1(30) in OI type-VIII osteoblasts. This observation is in line with the upregulated matrix-related genes in OI type-VIII MSCs. Supplement Figure 2 shows the comparison of gene expression between OI type-VIII and type-I MSCs and osteoblasts.

### Immune cell populations and their gene expression in OI patients

The immune cell composition in the bone marrow of OI type-I patients showed large variation per donor and exhibited marked deviations compared to healthy individuals (Figure 5A). In general, bone marrow T cells showed a decline in proportions from an average of 16.53% in healthy to an average of 6.29% in OI type-I, and 8,74% in OI type-VIII (Figure 5B). However, when looking specifically at CD4+ T cells in OI type-VIII, a large increase to 26.76% was observed (Figure 5B). Furthermore, monocyte subsets, especially CD16- classical monocytes, showed a drastic increase, from 1.78% to 22.66% in OI type-I and 8.69% in OI type-VIII (Figure 5B).

**Figure 5:**
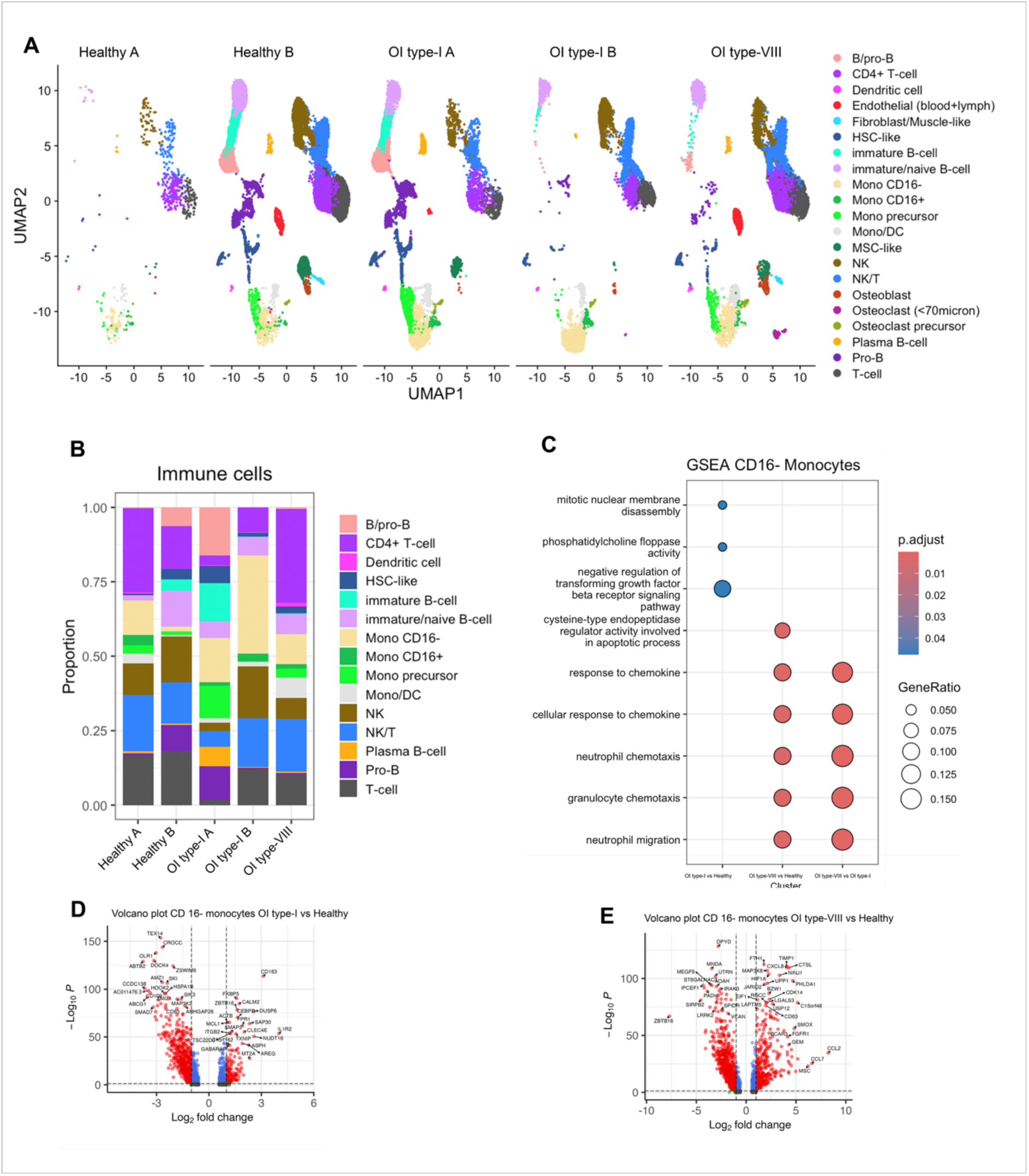
Immune cell composition and monocyte gene expression in OI patients. **(A)** UMAP plots showing immune cell distributions. **(B)** Bar plot of immune cell proportions in healthy controls (A and B), OI type-I (OI A and B), and OI type-VIII patients. **(C)** Gene Set Enrichment Analysis (GSEA) results for CD16- monocytes comparing OI type-I and OI type- VIII patients to healthy controls. **(D,E)** Volcano plots of differentially expressed genes in CD16- monocytes from OI type-I (D) and OI type-VIII (E) patients compared to healthy controls.

Further differential gene expression and GSEA analysis was performed in CD16- classical monocytes, as they are potential precursors of osteoclasts(31–33), and showed a large deviation between healthy and OI bone marrow. GSEA in CD16- classical monocytes revealed that pathways related to cell migration and chemotaxis were altered in the CD16- monocyte subsets in the bone marrow of OI patients (Figure 5C). Additionally, the negative regulation of TGF-β Receptor Signaling pathway was altered in CD16-monocytes of OI patients, which could be linked to immune dysregulation or bone remodeling. This observation was confirmed in the differential gene expression of OI type-I CD16- monocytes, where two negative regulators of TGF-β signaling(34): SMAD7 and CD109, are downregulated, potentially inducing excessive TGF-β signaling, which is a common mechanism in OI(35) (Figure 5D). In addition, upregulation of inflammatory genes(36–39), such as CD163, CEBPD, and IL1R2, suggests enhanced monocyte activation state compared to healthy controls. In OI type-VIII, CXCL8, CCL2 and CCL7 were upregulated, which are genes related to promoting inflammation and recruiting monocytes(40, 41) (Figure 5E). In addition, CD63, a marker may indicate a subset of classical monocytes primed for fusion into multinucleated cells(42), like osteoclasts, is upregulated. In addition, upregulation of HIF1A(43), potentially indicates a pathway for inducing osteoclast differentiation in these monocytes by enhancing RANKL induction(43). These findings in type-VIII CD16- monocytes align with the increased number of osteoclast and osteoclast precursors observed in this patient.

## Discussion

In this study, we explored whether changes in the bone marrow microenvironment may contribute to the variability seen in OI using single-cell RNA sequencing of patient-derived bone tissue. Our main finding indicates that, beyond the known collagen defects, OI bone shows reduced MSC numbers and lower expression of osteogenic genes in MSCs. Given the small sample size, our findings should be interpreted with caution. Still, the observed differences in both mesenchymal and immune cell populations point to a potentially altered bone remodeling environment in OI that may differ per patient.

Across both OI types MSC numbers were reduced, and key genes involved in osteogenic differentiation: RUNX1 and BMP5, were downregulated. Runx1 knockout models show severe defects in bone formation with increased adipocyte accumulation(44), while a notable phenotype in *Bmp5* mutant mice is the reduction or absence of the deltoid crest, a bony ridge formed by mechanical interaction between muscle and bone(45). In addition, previous studies(46) in OI MSCs have also reported reduced osteogenic differentiation alongside a shift toward adipogenesis, which is in line with our findings. Similarly, a previous study (47)of impaired pediatric bone development showed disrupted osteogenic differentiation accompanied by compromised bone architecture and changes across multiple bone marrow cell populations. Together, these data support the idea that impaired commitment toward the osteoblast lineage may contribute to reduced bone formation in OI, in addition to the underlying collagen defect. Whether these changes are a direct consequence of the collagen mutation or reflect adaptations to an altered bone microenvironment remains unclear. In addition to downregulated RUNX1 and BMP5, MSCs in OI type-VIII showed higher expression of matrix-related genes, including COL1A1, THBS2, and TIMP1, which is counterintuitive but may reflect a compensation mechanism for the defective collagen. In another study, although the early osteogenic marker (COL1A1) was downregulated, late matrix markers such as IBSP (osteosialic acid) were significantly elevated in osteoblasts from OI patients during in vitro differentiation compared with the control group (48). Together, these findings indicate that MSCs may be altered subtype-specific in OI, although these observations remain exploratory given the limited sample size.

This study showed that OI type-I was characterized by reduced osteoblast numbers, and little osteoclasts were detected. In contrast, in OI type-VIII, osteoblast numbers were increased and showed increased expression of several extracellular matrix genes (similar to the OI type-VIII MSCs). In addition, OI type-VIII bone showed an increase in osteoclasts, indicating a shift toward enhanced bone resorption and turnover. We cannot exclude osteoclast numbers may be influenced by bisphosphonate treatment, even though all patients received bisphosphonates, the timing and dosing regimens may have varied. Nevertheless, previous studies report substantial variability in osteoclast number and activity across OI cohorts and animal models, with significant heterogeneity observed between studies and modulation by age, disease severity, and underlying genetic mutations (49). Suggesting that osteoclast involvement in OI is not uniform, in line with our results. Individual osteoclast variability may also help explain the mixed clinical response to bisphosphonates(50), in addition to factors like patients age, genetic mutation, timing and the specific drug used. Thus, differences in osteoblast/osteoclast bone cell dynamics between patients may explain parts of variation in disease severity, and could provide direction when choosing the optimal therapy for individual patients.

Furthermore, our results indicate (longitudinal) bone growth may be affected via other mechanisms than impaired collagen formation. We show that growth hormone receptor was downregulated in osteoblasts of both OI types, and in OI type-VIII MSCs. This suggest impaired responsiveness to growth-related signals. Indeed, previous studies observed an increase in longitudinal bone growth and bone quality upon growth hormone treatment in OI patients (51, 52). In addition, the reduction of MSC-like cells we observed in OI patients, may limit the pool of osteoprogenitors that normally support growth plate progression, via both paracrine and matrix cues(53).Several studies demonstrate that disruptions in progenitor function or growth plate integrity can delay hypertrophic chondrocyte maturation and impair endochondral ossification in OI(54, 55). At the same time, expression of BMP5 was lower in OI MSCs, which is a key morphogen regulating chondrocyte proliferation and hypertrophy(56), and BMP5 has been directly implicated in growth-plate development(56). Together, this suggests that growth hormone and growth plate activity may play a role in pediatric OI, but the underlying cellular mechanisms at the level of the growth plate remain largely unexplored.

Changes in the bone microenvironment in OI are likely to affect not only bone-forming cells but also immune populations. Indeed, we observed a marked increase in CD16⁻ monocytes in OI bone marrow, accompanied by increased inflammatory and activated state in both OI types. Previous studies in patients also suggest heightened inflammatory processes in OI, such as one in 71 children with moderate to severe OI where elevated platelet counts were observed(57). In addition, a study using whole-blood RNA sequencing in OI types I-VII demonstrated dysregulation of inflammatory pathways and altered bone metabolic activity(58). Another study reported that the OIM (B6C3Fe a/a-Col1a2^oim^/J) murine model (with severe OI) showed splenomegaly and expanded CD11b+ myeloid populations, with elevated splenic osteoclast precursors, a pattern indicative of chronic inflammation and systemic myeloid dysregulation(59). Further evidence that inflammation may play a role in OI, was shown in a recent OI mice model(60), which reported fewer circulating regulatory T cells (Tregs) and higher inflammation, which disturbed bone remoddeling and increased bone loss and fragility. Adding back Tregs reduced inflammation, boosted osteoblasts, reduced osteoclasts, and improved bone structure. This indicates the immune profile of OI patients may represent a novel therapeutic target, with strategies aimed at reducing inflammation possibly alleviating symptoms in some OI patients.

This study has several limitations. First, the small sample size, including only three OI patients, limits the generalizability of our findings. In addition, scRNA-seq provides a snapshot of gene expression and does not capture dynamic processes such as lineage progression or disease development over time, nor does it allow conclusions about causality. Our analysis was restricted to bone and bone marrow-derived cells, and therefore does not account for systemic factors such as endocrine or metabolic signals that are known to influence bone homeostasis. Furthermore, while we observed changes in pathways related to growth plate regulation, we did not directly assess growth plate chondrocytes, limiting conclusions on cartilage-to-bone transition. Finally, the enzymatic dissociation required for single-cell analysis may introduce technical bias, including loss of certain cell types such as osteocytes and the induction of stress-related gene expression. Despite these limitations, this study provides a first single-cell view of the pediatric human OI bone microenvironment compared to healthy controls and offers a basis for future, more comprehensive studies.

In summary, this study provides initial insight into how differences in the bone marrow microenvironment may contribute to the heterogeneity seen in OI. We observed distinct cellular and molecular patterns between OI type I and type VIII, suggesting that the effects of collagen (OI type-I) and LEPRE1 (OI type-VIII) mutations extend beyond the collagen matrix itself and involve broader interactions with bone and immune cells. Our findings add to the hypothesis that disease variability in OI may not be explained by genetic defects alone, but also by how these defects shape the local cellular environment. While exploratory, this may help explain differences in disease severity and treatment response between patients. Future studies in larger cohorts are needed to further define these interactions and assess whether they can inform more targeted, patient-specific therapies.

## Data Availability

All data produced in the present study are available upon reasonable request to the authors. Upon publication in a peer-reviewed journal, all data will be made available online.

## Acknowledgements

The authors Nijhuis, Sakkers and Spaans are members of the European Reference Network on Rare Bone Diseases (ERN BOND).

---

**Supplementary Figure 1.**
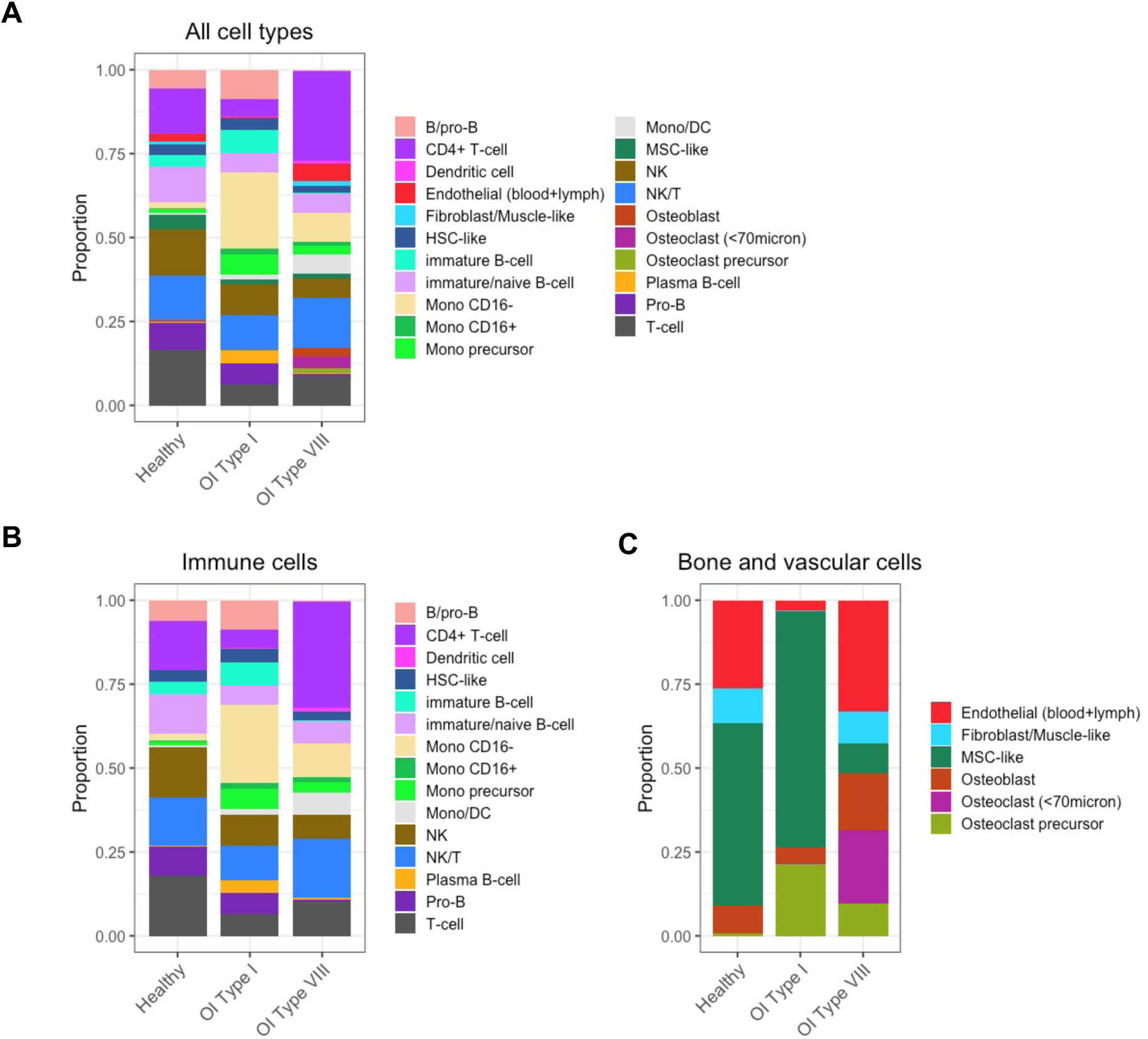
Proportional Changes in All Cell Types, Immune Cells, and Bone-Associated Cells Across OI Subtypes and Healthy Controls A Proportions of all detected cell types in healthy controls, OI type-I, and OI type-VIII. B Proportions of immune cells. C Proportions of bone and vascular cells.

**Supplementary Figure 2.**
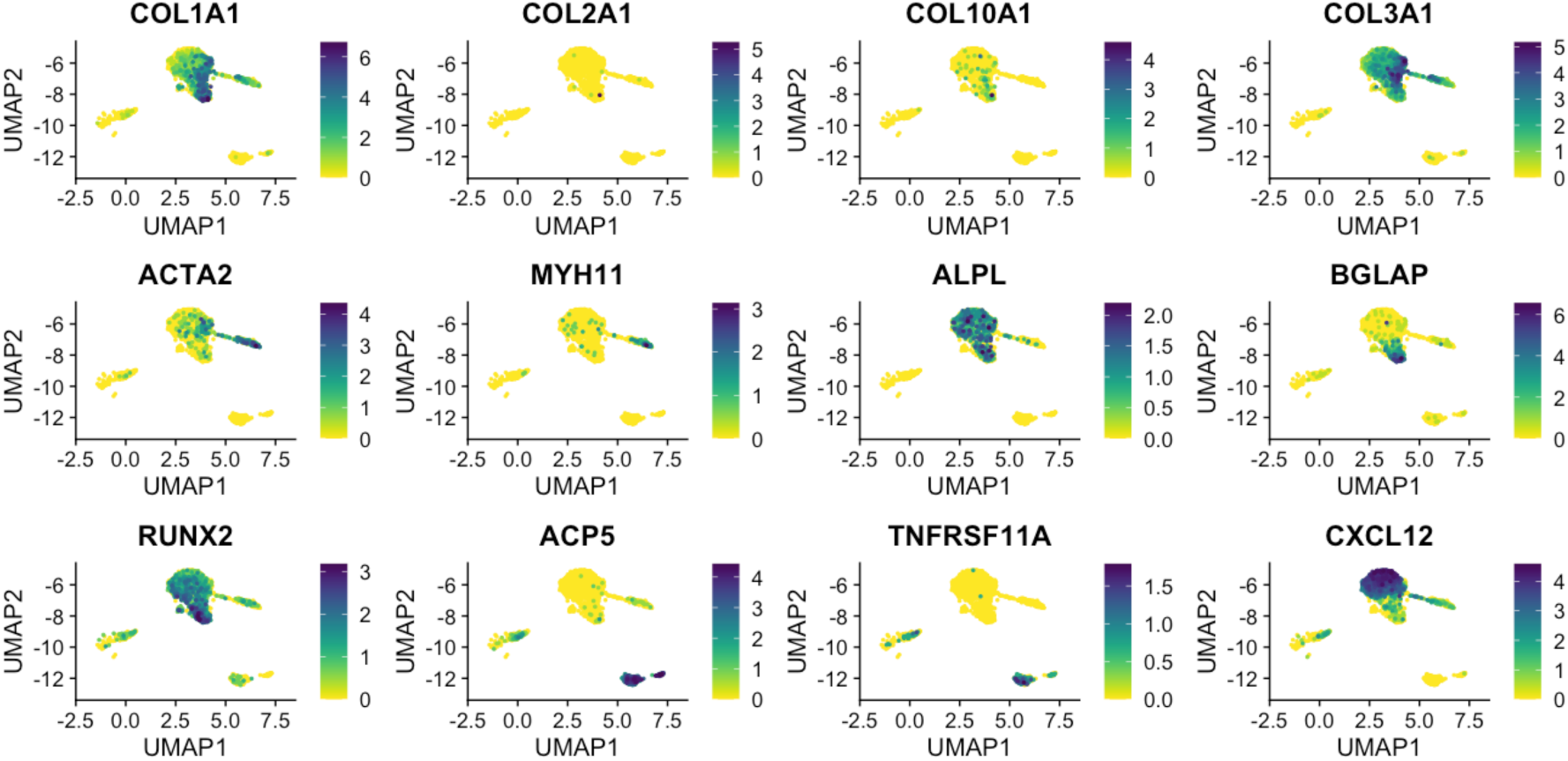
UMAP visualization of key bone-related gene expression across bone cell clusters of healthy individuals and OI patients. Expression levels are represented by a color gradient from yellow (low or no expression) to green (high expression). The distribution of bone-related cell types, including MSC-like, fibroblast/muscle-like cells, osteoblasts, osteoclasts, and osteoclast precursors, is annotated in Figure 3A.

**Supplementary Figure 3.**
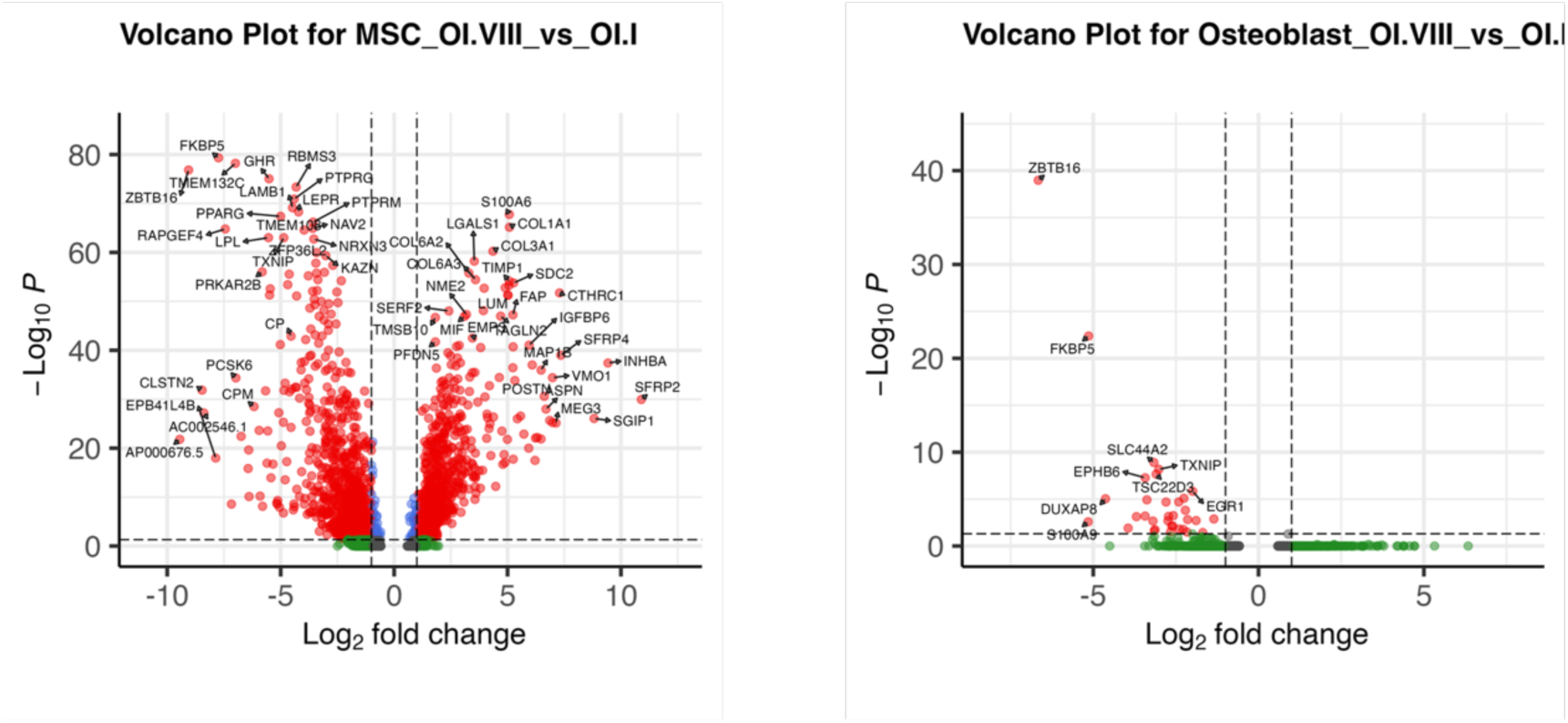
Volcano plots depicting differentially expressed genes in MSCs and osteoblasts.

